# Automated objective dystonia identification using smartphone-quality gait videos acquired in clinic

**DOI:** 10.1101/2020.06.09.20116954

**Authors:** Hanyang Miao, Keisuke Ueda, Toni S. Pearson, Bhooma R. Aravamuthan

**Author notes:** Correspondence to: B.R. Aravamuthan, Department of Neurology, Division of Pediatric Neurology, Washington University School of Medicine, 660 South Euclid Avenue, Campus Box 8111, St. Louis, MO, 63110-1093, USA.

## Abstract

**Background:** Dystonia diagnosis is subjective and often difficult, particularly when co-morbid with spasticity as occurs in cerebral palsy.

**Objective:** To develop an objective clinical screening method for dystonia

**Methods:** We analyzed 30 gait videos (640×360 pixel resolution, 30 frames/second) of subjects with spastic cerebral palsy acquired during routine clinic visits. Dystonia was identified by consensus of three movement disorders specialists (15 videos with and 15 without dystonia). Limb position was calculated using deep neural network-guided pose estimation (DeepLabCut) to determine inter-knee distance variance, foot angle variance, and median foot angle difference between limbs.

**Results:** All gait variables were significant predictors of dystonia. An inter-knee distance variance greater than 14 pixels together with a median foot angle difference greater than 10 degrees yielded 93% sensitivity and specificity for dystonia.

**Conclusions:** Open-source automated video gait analysis can identify features of expert-identified dystonia. Methods like this could help clinically screen for dystonia.

## Introduction

The most common cause of dystonia in childhood is neonatal brain injury resulting in cerebral palsy (CP).(1) In this etiological context alone, dystonia affects at least 0.5-1 per every 1000 live births in the United States.(1-3) Dystonia is difficult to identify in those with CP and is often confused for co-morbid spasticity.(4,5) This has led to a dystonia diagnostic delay of years in children with CP, which is often a longer diagnostic delay than for other etiologies.(5) Possibly contributing to this delay is that the gold standard for dystonia diagnosis remains subjective assessment by a motor phenotyping expert. Given that dystonia is characterized by its variability over time and may appear differently based on the type of voluntary movement trigger, objective criteria for dystonia diagnosis have remained elusive.(6) This may be particularly true for CP since different types and extents of brain injury could result in different manifestations of dystonia.(7)

Since dystonia can be identified visually, numeric readouts of limb movement trajectories have the potential to provide an objective method for dystonia diagnosis. Objective motor quantification techniques, like conventional video-based gait analysis, can give such readouts. However, conventional gait analysis measures have been unable to differentiate between dystonia and spasticity.(8) Commercially available systems are not easily adaptable to query the characteristics of gait that motor phenotyping specialists may commonly associate with dystonia. Furthermore, conventional video-based gait analysis systems are expensive and have a large physical footprint, preventing their widespread use at many clinical centers.

To address these issues, we sought to determine a clinically-feasible and objective method to aid in dystonia identification in children and young adults with CP during routine clinic visits. We hypothesized that quantification of movement trajectories using open-source deep neural network transfer learning software (DeepLabCut(9,10)) could be used to objectively identify the movement features cited by phenotyping experts as they differentiate between those with and without dystonia.

## Methods

This study was granted Human Subjects Research approval from the Washington University School of Medicine Institutional Review Board.

Gait videos of children and young adults diagnosed with CP and spasticity were reviewed for dystonia by three fellowship-trained pediatric movement disorders specialists (B.R.A., K.U., T.S.P.). These gait videos were all retrospectively selected from a database of videos recorded during routine outpatient clinic visits in the St. Louis Children’s Hospital Cerebral Palsy Center. Videos were recorded using an Apple iPad A1432 at 640×360 pixel resolution and 30 frames/second. Of note, this is the typical resolution and frames/second capability of standard definition video recording on most commercially available smartphones. Videos consisted of subjects walking barefoot in a straight line down a 15 foot hallway towards the camera. Subjects were all independently ambulatory but were allowed to use hand-held walking aids in the videos. Given that some subjects were holding onto walking aids in the videos while others were not, experts were only asked to assess the presence or absence of dystonia in the lower extremities.

Only videos in which the subject was unanimously deemed to display or not display lower extremity dystonia by all three specialists were used for further analysis. For this retrospective case-control study, 15 subjects whose gait videos displayed dystonia were compared to 15 subjects with videos that did not display dystonia. Subjects were chosen to have comparable etiologies of their CP: all had periventricular leukomalacia on their MRIs without history of intracranial surgery, brain malformations, genetic disease, or metabolic disease. Subjects with and without dystonia were of comparable age, sex, and Gross Motor Function Classification System Level(11) distribution (Table 1).

**Table 1.** Subject Clinical Details

|  | No Dystonia (n=15) | Dystonia (n=15) | p (t-test or Chi-square) |
| --- | --- | --- | --- |
| Gestational age at birth (weeks) | 29 (27-30) | 31 (29-33) | 0.10 |
| Sex (% male) | 53 (28-79) | 40 (15-65) | 0.46 |
| GMFCS (%) |  |  |  |
| I | 33 (9-57) | 13 (0-31) | 0.40 |
| II | 47 (21-72) | 53 (28-79) |  |
| III | 20 (0-40) | 33 (9-57) |  |
| Age in video (years) | 11.0 (9.6-13.3) | 12.9 (10.3-15.4) | 0.28 |

GMFCS – Gross motor function classifications system, p values are based on t-tests for gestational age and age in the video, and Chi-square tests otherwise

Quantitative automated motion tracking was done across all videos using open-source software (DeepLabCut). DeepLabCut (version 2.1.1(9,10)) utilizes deep neural network transfer learning methods to label user-defined nodes on any video and subsequently generate numerical readouts of limb movements without the application of markers or use of specialized cameras, even in sub-optimal backgrounds or lighting environments. DeepLabCut’s numerical readouts can be easily adapted to generate user-defined movement parameters. It has been applied to characterize movements as diverse as mouse pupil constriction to electric fish swimming to human hand reaching.(9,10) Given its broad applicability, this software allows for optimal analysis of patient videos obtained during routine clinical care, which are likely to be obtained in diverse settings and lighting conditions and under time-constraints that prohibit marker application. The neural network is trained on a relatively small number of user-labeled video frames (50-200 frames or 2-7 seconds of video) and can subsequently be used to automatically label all frames of any video acquired in comparable settings (e.g. the same clinic hallway). Output includes the X and Y coordinates of each user-defined node in each frame together with the likelihood of correct labeling (p-cutoff ranging from 0 to 1, with 1 indicating absolute certainty of labeling accuracy). We labeled 10 frames per video yielding 300 frames for training and testing the network (95% for training, 5% for testing). Frames were labeled while blinded to the identity of the patient or whether they had been identified as displaying dystonia in the video. A ResNet-50-based neural network was used for 500,000 training iterations to achieve a train error less than 2 pixels and a test error less than 3 pixels. We used a p-cutoff of 0.9 to condition the X,Y coordinates for further analysis.

Using the trained network, the following nodes were labeled in automated fashion and therefore agnostic to whether the video was identified to contain dystonia or not: bilateral midpoints of the patellae, midpoints between the lateral and medial malleoli, and third toes. Using the generated coordinates of these nodes, we determined three gait variables: variance of the inter-knee distance normalized to the maximum knee to ankle distance in the same frame, foot angle variance (larger of the left and right foot angle variances), and median difference of the foot angle between limbs (Figure 1A). These gait variables were chosen *a priori* based on characterizations of classic dystonic movements and postures(12,13) and on the consensus definition of dystonia which notes that variability and a voluntary movement trigger are key defining features.(6) Although determining the bilateral leg angles relative to the hips (iliac crests) would be the ideal measure to assess leg adduction, the hips were not reliably visible in these subject videos that were retrospectively chosen from videos acquired during routine clinical care. Therefore, inter-knee distance variance was examined in lieu of comparing leg angles between limbs and over time, though we were able to compute these measures for the feet.

**Figure 1.**
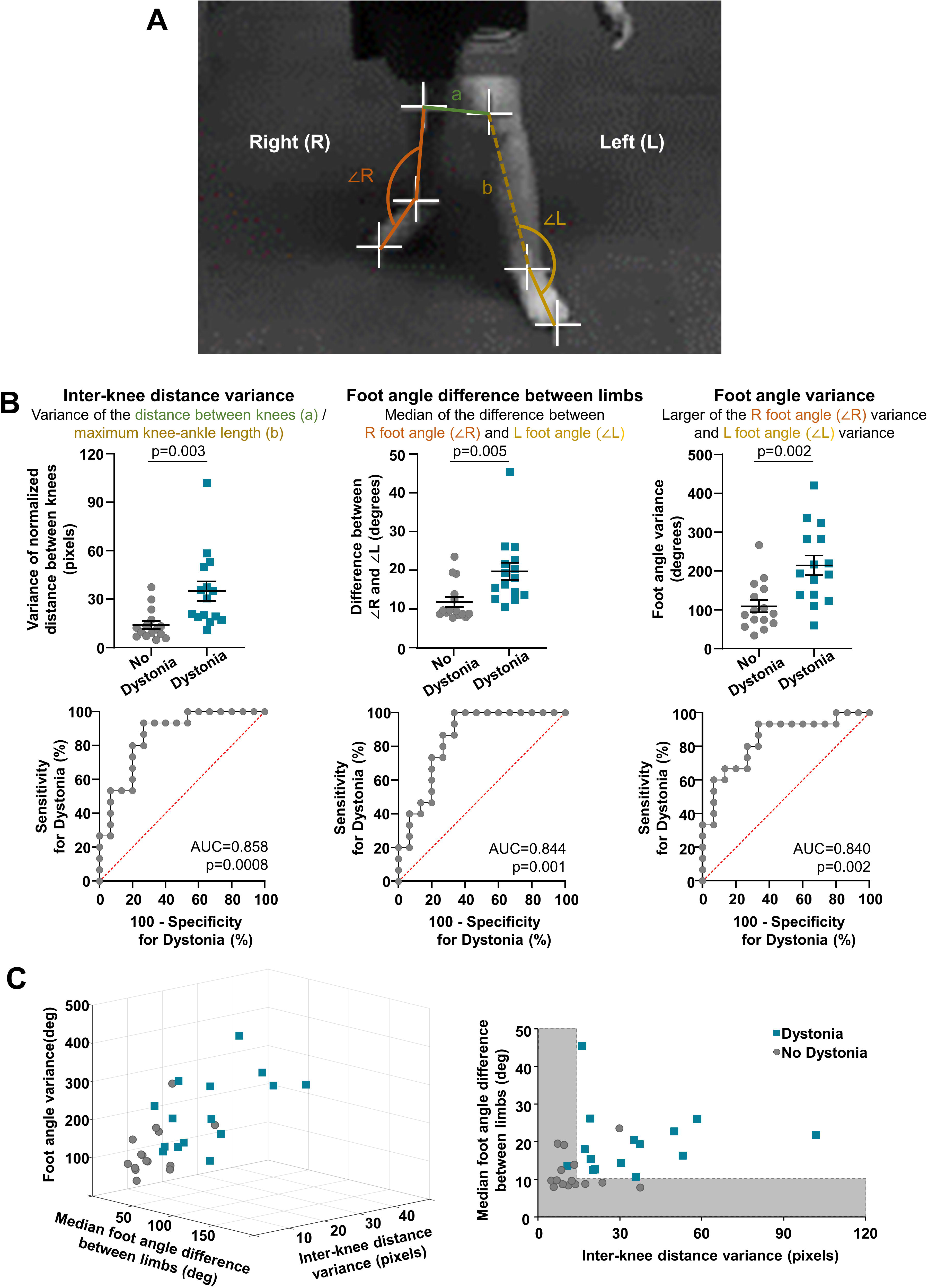
Calculation of gait variables from following automated limb labeling. A) Example of marker positions and features used to calculate gait variables in a single video frame. B) Comparison of gait variables between subjects who did or did not display dystonia in their gait videos using post-hoc Tukey HSD following MANOVA (top). Receiver operator characteristic areas under the curve were used to determine if these variables were significant predictors of expert-identified dystonia in videos (bottom). C) Segregation of subjects that did or did not display dystonia across all three variables (left). Inter-knee distance variance and median foot angle difference between limbs, when considered together provided the best separation between groups (right). Gray shaded areas indicate upper limit cutoff values for these two variables that could help distinguish between subjects that did or did not display dystonia in gait videos.

Data were first analyzed using MANOVA to assess whether the three dependent gait variables were different between videos that either did or did not display dystonia. MANOVA Pillai’s trace p<0.05 was deemed significant before employing post-hoc comparisons via Tukey’s HSD with a significance cutoff of p<0.05 (SPSS, IBM, Armonk, NY). Receiver operator characteristic (ROC) curves were used to determine the sensitivity and specificity of gait variables for predicting expert-identified dystonia (Graph Pad Prism 8, GraphPad Software, San Diego, CA).

## Results

The assessed gait variables were significantly different between videos that did display expert-identified dystonia compared to those that did not (MANOVA, Pillai’s trace F=6.336, p=0.002). Inter-knee distance variance over time (p=0.003), foot angle variance over time (p=0.005), and the median difference of the foot angle between limbs (p=0.002) were all significantly greater in videos that displayed expert-identified dystonia. ROC curves of these three gait variables revealed that all were significant predictors of expert-identified dystonia. In isolation, all variables offered comparable sensitivity and specificity for dystonia prediction. Inter-knee distance variance greater than 18 pixels was 80% sensitive and 80% specific for dystonia with ROC area under the curve (AUC) of 0.858 (95% CI 0.722-0.994, p=0.0008). Median difference of the foot angle between limbs greater than 14 degrees was 73% sensitive and 80% specific for dystonia (AUC 0.844, 95% CI 0.698-0.991, p=0.001). Foot angle variance greater than 136 degrees was 80% sensitive and 73% specific for dystonia (AUC 0.840, 95% CI 0.695-0.985, p=0.002) (Figure 1B). When considering these variables together, inter-knee distance variance greater than 14 pixels together with a median difference of the foot angle between limbs greater than 10 degrees was 93% sensitive and 93% specific for dystonia (Figure 1C). Additional consideration of foot angle variance did not improve the sensitivity or specificity for dystonia.

## Discussion

Quantifiable gait variables can distinguish between subjects with CP who do or do not display lower extremity gait dystonia using only smartphone-quality gait videos obtained during routine clinic visits. These variables were able to identify features of expert-identified dystonia with high specificity and sensitivity.

These gait variables are representative of the consensus definition of dystonia, quantifying movement features that are variable over time and between limbs.(6) They can be quantified from videos taken using smartphones or tablet computers in an outpatient clinic without the use of specialized cameras or markers. Video processing can be done with open-source software.(9,10) Therefore, the methods outlined here are broadly applicable in a cost-effective manner in outpatient clinic settings.

Regarding implementation, each clinical center would need to computationally train a neural network to distinguish patient body parts in a representative set of videos filmed in their local clinic environments. Training can take several days of processing time with a traditional personal computer (central processing unit), but can occur within 48 hours with a graphics processing unit. After the network is trained, videos acquired in comparable clinic environments can be labeled within minutes depending on the length of the video. Therefore, after the initial requirement for neural network training, video analysis can happen within the same day of the clinic visit, or even during the visit itself.

It will be important to validate this method in large data sets of ambulatory patients with and without dystonia and to include subjects with gait dystonia from different etiologies. Comparable techniques could be used to determine which variables correlate with expert-identified dystonia in other parts of the body during other motor tasks. Therefore, the results described here could be the first in a long line of objective and quantifiable movement features that could help aid clinical dystonia identification. We ultimately envision that these techniques could help practitioners initially screen patients for features which may be consistent with dystonia, particularly at centers where dedicated movement disorders specialists may not be readily available.

## Data Availability

All data will be made available to qualified investigators upon request.

## Author roles

(1) Research project: A. Conception, B. Organization, C. Execution; (2) Statistical Analysis: A. Design, B. Execution, C. Review and Critique; (3) Manuscript Preparation: A. Writing of the first draft, B. Review and Critique

H.M. 1B, 1C, 2C, 3B

K.U. 1B, 1C, 1C, 3B

T.S.P. 1C, 2C, 3B

B.R.A 1A-C, 2A, 2B, 3A

